# Complex Regulation of Protocadherin Epigenetics on Aging-Related Brain Health

**DOI:** 10.1101/2024.04.21.24306143

**Authors:** Vanessa Schmithorst, Abha Bais, Daryaneh Badaly, Kylia Williams, George Gabriel, Rafael Ceschin, Julia Wallace, Vince Lee, Oscar Lopez, Annie Cohen, Lisa J. Martin, Cecilia Lo, Ashok Panigrahy

**Affiliations:** UPMC Children’s Hospital of Pittsburgh, University of Pittsburgh, Department of Radiology; University of Pittsburgh Department of Developmental Biology; Child Mind Institute; University of Pittsburgh Integrative Systems Biology; University of Pittsburgh Department of Neurology; University of Pittsburgh Department of Psychiatry; Department of Pediatrics Cincinnati Children’s Hospital Medical Center and the University of Cincinnati College of Medicine

## Abstract

Life expectancy continues to increase in the high-income world due to advances in medical care; however, quality of life declines with increasing age due to normal aging processes. Current research suggests that various aspects of aging are genetically modulated and thus may be slowed via genetic modification. Here, we show evidence for epigenetic modulation of the aging process in the brain from over 1800 individuals as part of the Framingham Heart Study. We investigated the methylation of genes in the protocadherin (PCDH) clusters, including the alpha (*PCHDA*), beta (*PCDHB*), and gamma (*PCDHG*) clusters. Reduced *PCDHG*, elevated *PCDHA*, and elevated *PCDHB* methylation levels were associated with substantial reductions in the rate of decline of regional white matter volume as well as certain cognitive skills, independent of overall accelerated or retarded aging as estimated by a DNA clock. These results are likely due to the different effects of the expression of genes in the alpha, beta, and gamma PCHD clusters and suggest that experience-based aging processes related to a decline in regional brain volume and select cognitive skills may be slowed via targeted epigenetic modifications.

## Introduction

The protocadherin (PCDH) gene clusters comprising three distinct gene families (α, β and γ) encode homotypic cell adhesion proteins regulating synaptic connectivity via specification of neuronal identity with combinatorial expression of genes in the three clusters ^1–3^. Studies in mice show that deficiency of PCDH gene clusters results in perturbed neural connectivity and behavioral deficits, impacting functional synaptic connections^3–5^. Here, we investigated the relevance of methylation in the PCDH gene clusters to aging-related regional brain volume loss and domain-specific neurocognitive outcomes leveraging data collected from the well-phenotyped Framingham Heart Study offspring cohort comprising of 1800 subjects for which genome-wide DNA methylation data, brain MRI, and longitudinal cognitive assessments are available. Reduced *PCDHG*, elevated *PCDHA*, and elevated *PCDHB* methylation levels were associated with substantial reductions in the rate of decline of regional white matter volume as well as certain cognitive skills, independent of overall accelerated or retarded aging as estimated by a DNA clock.

In contrast to overall DNA methylation which decreases with age, the PCDH gene clusters show striking increase in DNA methylation with age and are referred to as an aging DNA methylated region (DMR), an aging-DMR ^6^. Epigenetic alterations are a hallmark of aging but whether changes in DNA methylation in the PCDH clusters may impact human white matter structure and domain-specific neurocognitive outcomes is currently unknown ^7^.Proteins expressed via the protocadherin (PCDH) gene clusters have been shown to play an important role in cortical development, including neuronal differentiation and migration, axon outgrowth, dendritic arborization, and synaptogenesis^1^. However, their role in normal aging processes is less well known. Here, we investigate the role of methylation in the *PCDHA*, *PCDHB*, and *PCDHG* clusters in moderating brain volume loss and cognitive decline in a large cohort of normal middle-aged and older adults as part of the Framingham Heart Study (FHS).

## Material and Methods

### Participants

For the current study, we acquired subject data from the Framingham Heart Study (FHS) Offspring Cohort, which was formed in 1971 to include children of participants consented into the original study. Between 1999 and 2005, participants in the Offspring Cohort were recruited for ancillary magnetic resonance imaging (MRI) and neuropsychological testing (NPT) (*n* = 2433) and, between 2005 and 2008, a subset also participated in an epigenetic DNA methylation sub-study (*n* = 1936). Subjects missing any of the covariates, outcomes from brain MRI (*n* = 460), neuropsychological measures (*n* = 4), both MRI and neuropsychological measures (*n* = 2106), or DNA methylation studies (*n* = 497), were excluded from the study. FHS data were obtained through the database of Genotypes and Phenotypes (dbGAP, http://dbgap.ncbi.nlm.nih.gov;) The study was also internally approved by the University of Pittsburgh IRB - CR20010180-005).

### Imaging Outcomes

MR brain imaging outcomes were acquired using 1.0/1.5T scanners, and volumes were quantified from T2-weighted double spin-echo coronal sequences.^8^ Volumetric data (gray matter, white matter) was available from frontal, temporal, occipital, and parietal lobes. Participants’ earliest usable MRI exam was matched to the closest usable NPT. The mean difference between exam dates was 1.02 days, through most subjects (*n* = 1905) completed both study portions in one day.

### Neuropsychological Outcomes

Neuropsychological measures included: the Boston Naming Test (BNT30), a measure of language assessing confrontation naming; the Wechsler Memory Scale (WMS) Logical Memory Immediate Recall (LMI), Delayed Recall (LMD), and Recognition (LMR) and the WMS Verbal Paired Associates Total Immediate Recall (PASI) and Delayed Recall (PASD), measures of verbal memory; the WMS Visual Reproductions Immediate Recall (VRI), Delayed Recall (VRD), and Recognition (VRR), measures of visual memory; the Trail Making Test Part A (TRA) and Part B (TRB) measures involving visual search, motor speed, and executive functions; the Hooper Visual Organization Test (HVOT), a test of visuospatial skills; the Similarities subtest (SIM) from the Weschler Adult Intelligence Scale, Fourth Edition (WAIS-IV), a measure of verbal reasoning; and the Word Reading subtest from Wide Range Achievement Test (WRAT), a measure of acquired verbal skill often used to assess premorbid intelligence.

### DNA Methylation

At Exam 8, participants provided blood samples to be analyzed in genetic studies, including those examining DNA methylation levels, which were quantified using the Infinium HumanMethylation450 BeadChip platform from Illumina. DNA methylation data have been deposited and are available from the dbGaP web site, under phs000724.v8.p12.^9^ For the current study, we investigated total, PCDHA, PCDHB, and PCDHG methylation in relation to participant factors and outcomes. Raw idat files from two different consent cohorts were imported and processed separately in the R programing environment. Quality control, normalization and filtering were performed using the R packages *minfi* [doi: 10.1093/bioinformatics/btu049] and *sesame* [doi: 10.1093/nar/gky691]. Briefly, P-values were calculated using pOOBAH (P-value with out-of-and array hybridization), and noob (Norm-Exp deconvolution using out-of-band probes) background correction and non-linear dye bias correction was performed. Samples with >=100,000 failed probes were filtered as well as those where predicted sex did not match reported sex. Probes with P-value >=0.05 as well as low-quality probes (as implemented in *sesame*) were masked. All probes with missing values in >20% of samples, probes on sex chromosomes, cross-reactive probes [<u>10.4161/epi.23470</u>] as well as probes associated with SNPs were excluded. Data from both labs were pooled together yielding a dataset with 397, 436 sites and 2,667 samples (489 from lab#1 and 2,178 from lab#2). DNA methylation age (DNAmAge) as well as phenological age (phenoAge) were calculated using the ENMix R package [<u>10.1093/nar/gkv907</u>].

### Statistics

To examine the relationship between age, methylation, and MRI and NPT outcomes, we performed the following analyses. (For ease in interpretation, all methylation values (*PCDHA, PCDHB, PCDHG* and total) were normalized to unity standard deviation). The outcomes (dependent variables) were either brain volumes or cognitive performance. The best model for each outcome was found using the following procedure. All models included chronological age as an independent variable, and subject sex, packset used to estimate the methylation levels, age squared (to make any found interaction more robust) and accelerated age (ACCAGE) and age-X-ACCAGE interactions as covariates of no interest. ACCAGE and its interaction were included as covariates in order to better isolate the specific effects from each locus. Models were tested for each methylation value: 1) Completely excluded from the model; 2) Main effect only; 3) Main effect and interaction (with the independent variable); 4) Main effect and quadratic term; 5) Main effect, interaction, and quadratic term. As there are 5 options for each of four methylation values, a total of 5^4 = 625 linear regression models were tested. The best model was selected using the Aikake’s Information Criterion (AIC); values and standard errors of the parameters from the best model are displayed in Supplemental Tables 1-4. For selected cognitive tests and brain regions (Figures Y), age trajectories (with standard errors) were then plotted using the optimal models and regression parameters for given methylation values two standard deviations above and below the mean, to illustrate the difference in trajectories between individuals with high and low methylation values. We repeated these analyses with clinical risk factors (stroke, diagnosis of mild cognitive impairment/Alzheimer’s) as the outcomes using logit regressions and also with white matter hyperintensity volume as the independent variable (chronological age included as covariate of no interest).

## Results

We analyzed a subset of the FHS Offspring Cohort (Supplemental Figure 1), which demonstrates the following characteristics compared to the rest of the cohort: the patients excluded from the original Offspring Cohort were older, less educated, had increased brain injury and poorer performance on cognitive testing (Supplemental Table 1), suggesting that the sample included and analyzed in this study was relatively healthier and may be less confounded by the effect of brain injury. In the analyzed subset, DNA methylation in the protocadherin clusters increases with age, even as the overall DNA methylation genome-wide decreases (Supplemental Figure 2).

For each brain region and type (gray/white matter) except frontal gray, an increase in *PCDHA* methylation was associated with a retardation of age-related volume loss in both white matter (Figure 1A, Figure 1B, Figure 1D, Figure 1F, Supplemental Table 2) and grey matter (Figure 1C, Figure 1E, Figure 1G, Supplemental Table 2) of multiple lobes. A one-standard-deviation increase in *PCDHA* methylation resulted in a retardation of age-related regional brain volume loss by about one-fifth to one-third. An increase in *PCDHA* methylation also resulted in age-related preservation of acquired verbal skill as measured by Word Reading on the Wide Range Achievement Test (WRAT), often considered a “hold” test of premorbid skill (Figure 2A, Supplemental Table 3); however, it also resulted in an acceleration of cognitive loss (visual search, motor speed, and sequencing) as measured by the Trail Making Test Part A (TRA) (Supplemental Table 3). Increased *PCDHA* methylation also resulted in a greater effect of white matter pathologies (WM hyperintensity volume) on cognitive performance (Supplemental Table 4) for the Boston Naming Test (BNT30) (language function) and Similarities (SIM) from the Wechsler Adult Intelligence Scale, Fourth Edition (verbal reasoning).

**Figure 1:**
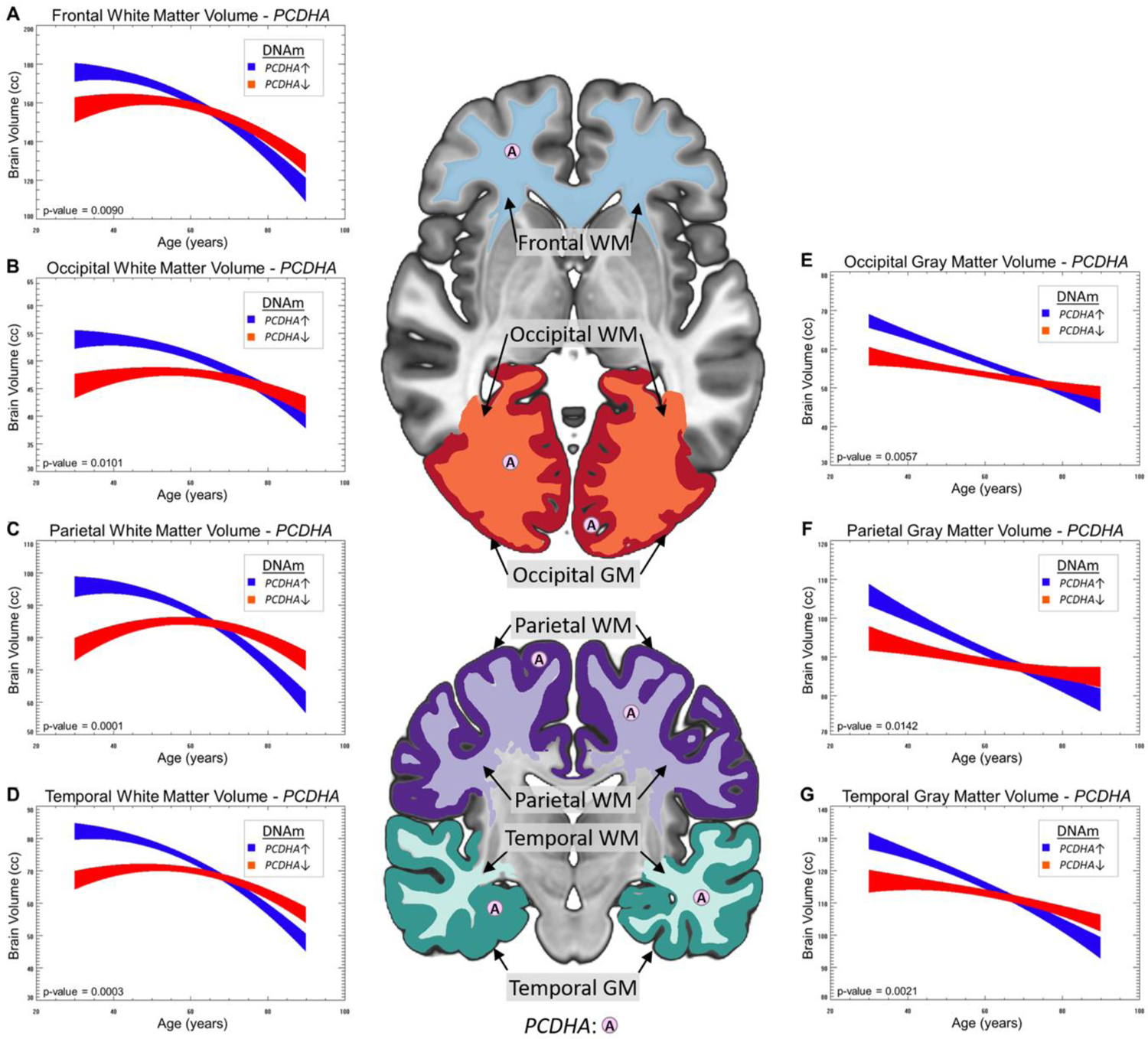
Association of Between PCDH Loci (alpha) and Total DNA Methylation Levels and Chronological Age-Related Decline in Regional Brain Volumes. For each brain region and type (gray/white matter) with the exception of frontal gray, an increase in *PCDHA* methylation resulted in a retardation of age-related volume loss in both white matter (Figure 1A, Figure 1B, Figure 1D, Figure 1F) and grey matter (Figure 1C, Figure 1E, Figure 1G) of multiple lobes. A one-standard-deviation increase in *PCDHA* methylation results in a retardation of age-related regional brain volume loss by about one-fifth to one-third.

**Figure 2:**
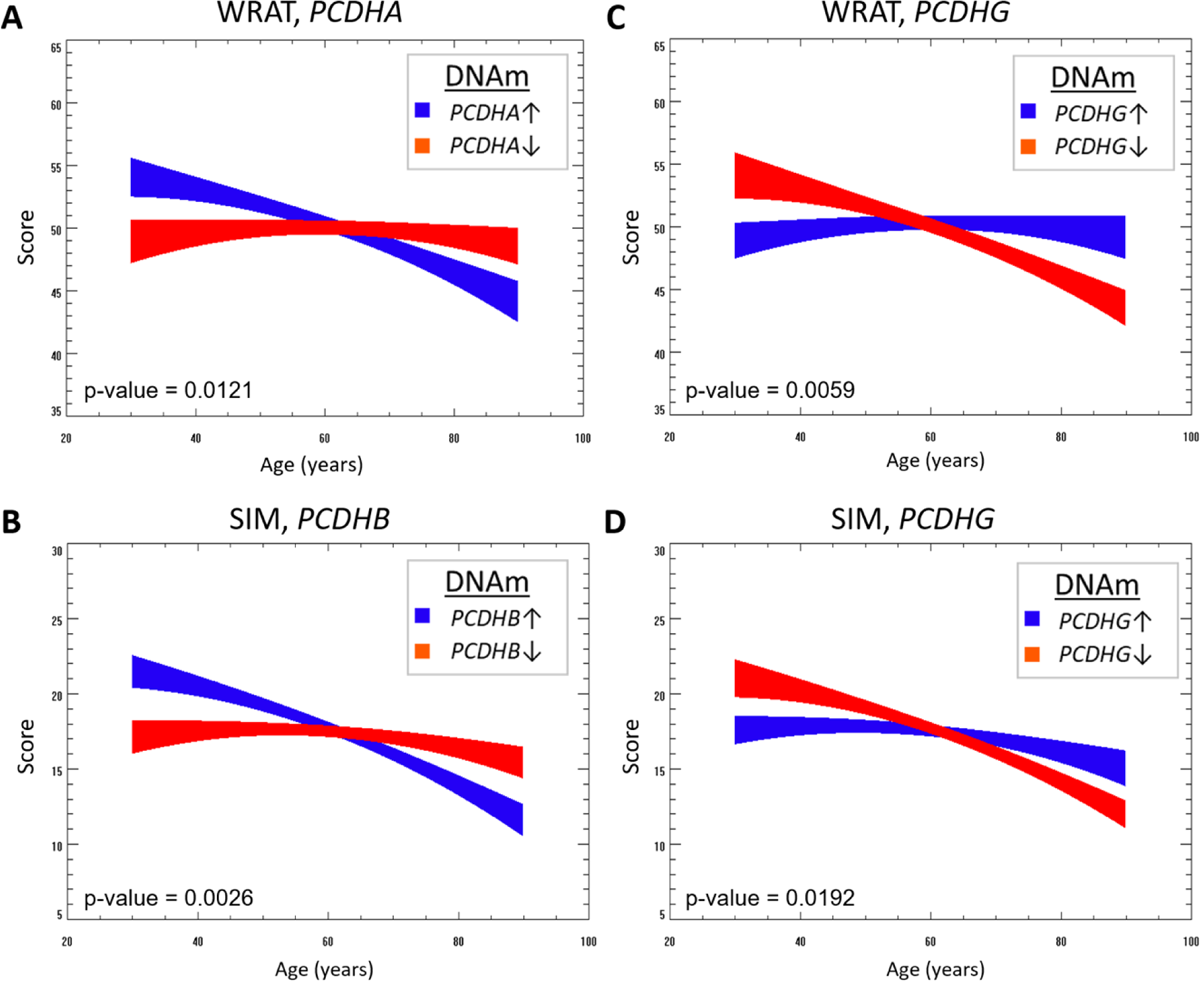
Association of Between PCDH Loci (alpha, beta, gamma) DNA Methylation Levels and Chronological Age-Related Decline in Neurocognitive Outcomes; An increase in *PCDHA* methylation results in age-related retardation of cognitive loss as measured by the WRAT (Figure 2A); Similarly, an increase in *PCDHB* methylation also resulted in retardation of age-related cognitive decline (Figure 2C; Supplemental Table 2) for the SIM. An increase in *PCDHG* methylation also was associated with an acceleration of age-related cognitive decline (Figure 2B, and Figure 2C; Supplemental Table 2) for the WRAT and the SIM.

An increase in *PCDHB* methylation resulted in a retardation of age-related volume loss in frontal gray matter (Figure 3; Supplemental Table 2) and a retardation of age-related cognitive decline (Figure 2B Supplemental Table 3) for the SIM (verbal reasoning). Increased *PCDHB* methylation also resulted in a smaller effect of white matter pathology (Supplemental Table 4) for the WRAT (acquired verbal skill).

**Figure 3:**
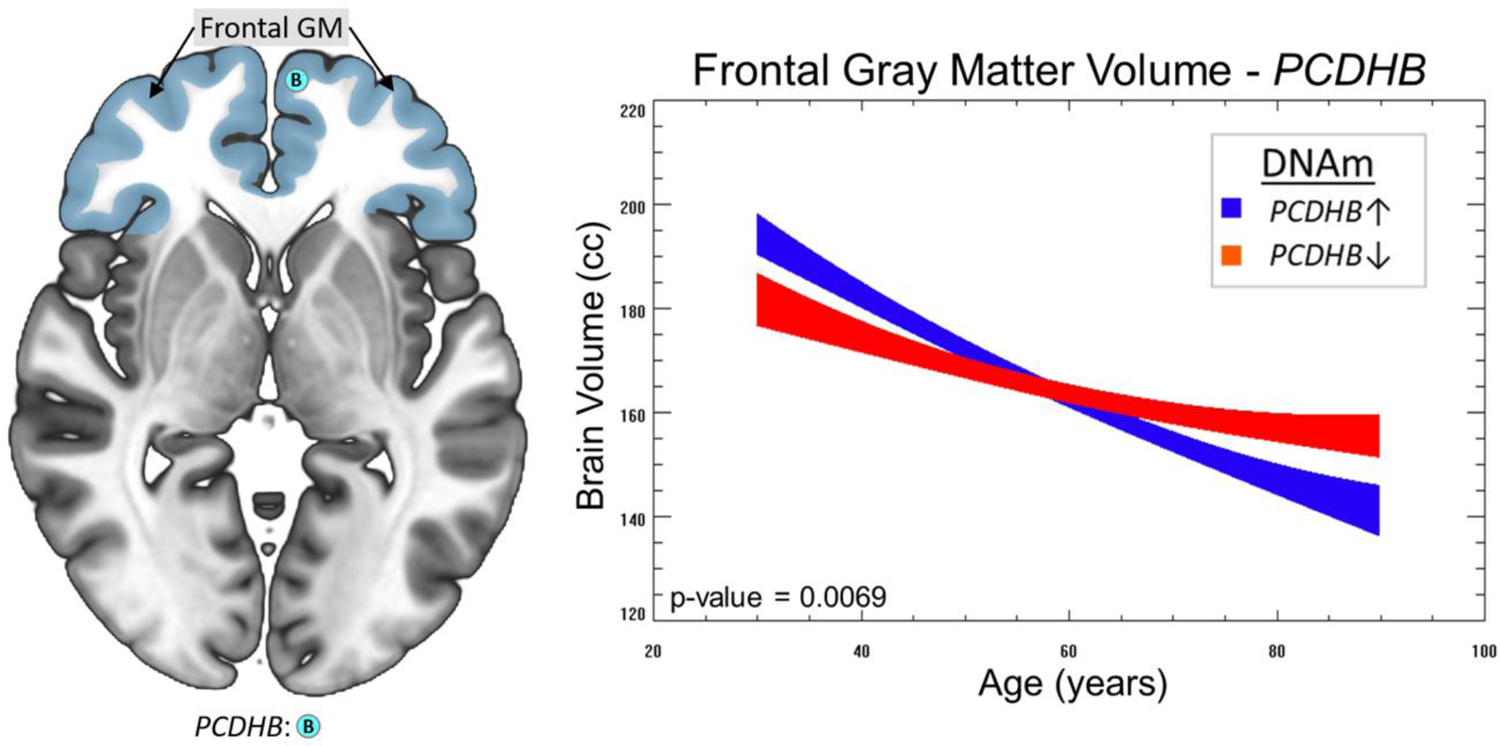
Association of Between PCDH Loci (beta,) and Total DNA Methylation Levels and Chronological Age-Related Decline in Regional Brain Volumes: An increase in *PCDHB* methylation resulted in a retardation of age-related volume loss in frontal gray matter.

Finally, an increase in *PCDHG* methylation resulted in an *acceleration* of age-related volume loss in parietal white matter (Figure 4A; Supplemental Table 2) and temporal white matter (Figure 4B, Supplemental Table 2). An increase in *PCDHG* methylation also was associated with an acceleration of age-related cognitive decline (Figure 2C, and Figure 2D Supplemental Table 3) for the WRAT (acquired verbal skill) and the SIM (verbal reasoning). Increased *PCDHG* methylation also resulted in a greater effect of white matter pathology (white matter hyperintensity volume) (Supplemental Table 4) for the WRAT (acquired verbal skill).

**Figure 4:**
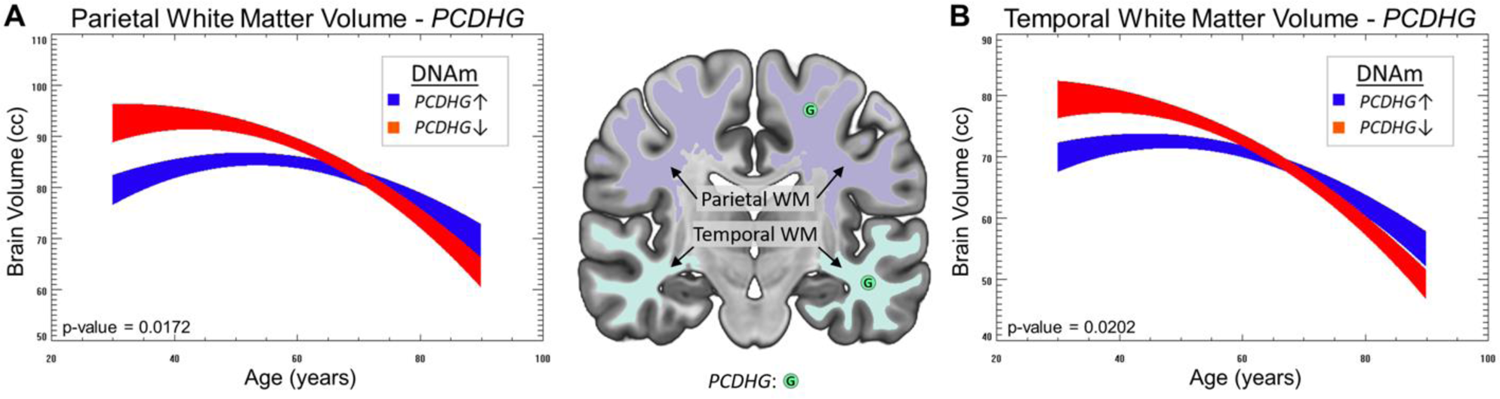
Association of Between PCDH Loci (gamma) and Total DNA Methylation Levels and Chronological Age-Related Decline in Regional Brain Volumes: Finally, an increase in *PCDHG* methylation resulted in an *acceleration* of age-related volume loss in parietal white matter (Figure 4A) and temporal white matter (Figure 4B).

While no age-X-methylation interactions were included as part of the optimal model for clinical risk (Supplemental Table 5), and the main effects included failed to reach significance (after family-wise error), the results about stroke merit some comment. The magnitude of the main effect for PCDHA methylation is three to four times the main effect for age (Supplemental Table 5), indicating that (if the ML estimates are the true values) a one standard deviation change in methylation values results in a greater stroke risk equivalent to three or four years of aging. The standard errors on these values are too large to reach statistical significance, but the possibility of elevated or reduced stroke risk due to *PCDHA* methylation warrants further investigation, as our results are limited by the small number of participants with the clinical condition (Stroke: N = 97; MCI: N = 123; Alzheimer’s: N = 53).

While not the main focus of our analysis, an increase in total methylation did not show any effect on brain volume loss; however, it was associated with an accelerated rate of cognitive decline (Supplemental Table 3) for Trail Making Test Part B (TRB) (executive function), resulting in a greater effect of white matter pathology (Supplemental Table 4) for TRB and a greater risk of MCI but a smaller risk of Alzheimer’s (Supplemental Table 5).

## Discussion

Our results show that normal aging processes affecting the brain and cognition are regulated by epigenetic configurations, particularly at *PCDH* loci. Interestingly, the A, B, G clusters affect aging processes in different ways, with the optimal configuration for aging appearing to differ from that during development. Unlike the characteristic pattern often seen for Alzheimer’s, in which memory impairments are often an initial and core feature, we showed that PCDH methylation affects a set of cognitive skills unrelated to memory; this is consistent with cognitive loss due to Alzheimer’s resulting from a different process than that resulting from normal aging. Finally, stroke risk and effect of white matter pathology on cognition are also affected by epigenetic configuration at the *PCDH* loci, indicating an effect on cognitive and vascular resiliency.

Our results clearly demonstrate that increased *PCDHA/PCDHB* methylation and decreased *PCDHG* methylation are associated with slower age-related rate of regional brain volume decline and slower age-related rate of cognitive decline for the specific abilities. As the level of DNA methylation in the PCDH cluster is correlated with gene expression in the PCDA cluster, our results suggest that increased expression of the *PCDHG* cluster and decreased expression of the *PCDHA/PCDHB* cluster may be protective during normal aging. ^7^ We hypothesize that these results reflect optimal conditions for synaptogenesis during aging, specifically synaptic specificity, which differ from optimal conditions during neurodevelopment. Our results are orthogonal to those found with total methylation, which did not affect brain volume loss, only showed an effect on tests largely tapping executive function (TRA/TRB), and were associated with MCI/Alzheimer’s risk but not stroke.

The role of protocadherins during neurodevelopment is well understood. cPcdhs are arranged in 3 gene clusters that encode 58 distinct isoforms of α-, β-, and γ-cPcdhs. ^10–12^ They impart neurons with single-cell identity by differential (and largely stochastic) expression of cPcdhs, which form combinatorial cPcdh recognition complexes.^13–15^ These molecules mediate crucial circuit formation functions^16^ including dendritic self-avoidance^17^, axonal tiling^18^, and cell survival. ^16,18–21^ Functional neural circuit construction requires a specific and organized regulation of cell-cell interactions in almost at all developmental stages, including neuronal differentiation, neuronal migration, axon outgrowth, dendrite arborization and synapse formation and stabilization.^22,23^ Cell-cell recognition through cell adhesion molecules is central to establishing this coordination; as cell-type specific surface molecules provide unique cellular surface identities and molecular diversity through their extracellular interactions that ultimately determine the formation of precise neural circuitry.^24,25^ Any mistake, error or mutation that leads to the formation of incorrect or altered neuronal connections can result in a number of neurodevelopmental disorders. The ability of neurites of individual neurons to distinguish between themselves and neurites from other neurons and to avoid self (self-avoidance) plays a key role in neural circuit assembly in both invertebrates and vertebrates. Similarly, when individual neurons of the same type project into receptive fields of the brain, they must avoid each other to maximize target coverage (tiling). Counterintuitively, these processes are driven by highly specific homophilic interactions between cell surface proteins that lead to neurite repulsion rather than adhesion. The clustered Pcdh proteins are essential for self-avoidance and tiling of neuronal processes as well as proper assembly of neuronal connectivity and cortical neuron migration.^17,18,26–29^ Their mutations or dysregulations are associated with numerous neuropsychiatric and neurodevelopmental disorders.^2,30^

However, during normal aging, the optimal situation is different, with the structural architecture of the brain already in place; therefore, there is much less risk of forming autapses or improper tiling. Brain volume loss due to normal aging (which does not involve neurodegeneration and neuronal loss) is distinct from that due to pathologies such as Alzheimer’s. Instead, during normal aging, the size and shape of neurons changes (including dendritic arbors), as does the axonal architecture (e.g., the distance between nodes of Ranvier increases with age in myelinated axons). Unlike abnormal aging from Alzheimer’s which is often characterized by a greater loss of memory function, normal aging may be related to loss of complex cognitive functions that tap, for instance, reasoning, speeded processing and executive functioning.

Therefore, synaptogenesis during normal aging is a likely mechanism for maintaining cognitive function in the presence of synaptic loss and efficiency, as well as retarding the process of synaptic loss. Thus, we hypothesize that synaptic specificity, critical to optimal developmental processes, may be harmful during normal aging. Our data shows that lower levels of *PCDHA/B* methylation, corresponding to higher levels of *PCDHA/B* expression, result in higher levels of brain volume and certain cognitive functions in early adulthood and middle age, associated with optimal brain development, suppressing the formation of autapses and promoting proper tiling. However, higher levels of *PCDHA/B* result in accelerated aging processes, with increased volume loss rates and select cognitive decline, which we hypothesize is related to the inhibition of synapse formation due to homophilic PCDH interactions.

The results for *PCDHA/B* are the direct opposite for *PCDHG,* in which *increased* methylation (corresponding to decreased expression) results in greater regional white matter volume and cognitive performance in certain domains in early adulthood/middle age but accelerated age-related decline. *PCDHG* is associated with dendritic arborization^31,32^ in which homophilic interactions *increase* dendritic arbor complexity (unlike the case for synaptogenesis in which homophilic interactions result in repulsion). Thus, lower levels of *PCDHG* expression likely accelerate aging-related dendritic arbor loss. Regarding development, *PCDHG* over-expression would likely result in over-complexity in dendritic arborization, impairing normal neurodevelopmental processes.

*PCDHG* under-expression (due to hypermethylation) would also result in neurodevelopmental impairment as has been seen in previous research^7^; however, our sample is mostly neurocognitively normal and thus would not include many individuals with *PCDHG* under-expression during development.

Our hypothesis of differential synaptogenesis dependent on PCDH methylation is consistent with the high degree of domain-specificity seen in the neurocognitive results. The rate of age-related decline on neurocognitive tasks which depend on language function is affected by PCDH methylation, while the same is not seen for executive function/working memory tasks. We would hypothesize that synaptogenesis enables aging individuals to retain more language ability in a manner similar to second language acquisition. Second language acquisition has been shown to be a factor in the development of “cognitive reserve” which delays cognitive decline in dementia patients^33^ and synaptogenesis is one of the major mechanisms involved. Also, the neurophysiology of second language acquisition has been shown to be quite similar to that of first language acquisition ^34^, further supporting our hypothesis. By contrast, working memory is thought to involve synaptic facilitation ^35^, possibly aided by astrocyte-mediated gliotransmission ^36^. More broadly speaking, executive function (comprising working memory, inhibition/cognitive control, and set shifting/cognitive flexibility, among other skills) is, during development, associated with a refinement process involving increasing specialization of the prefrontal cortex ^37^ and synaptic pruning combined with greater activation in existing synapses ^38^. Thus, synaptogenesis would be expected to provide little benefit regarding age-related cognitive decline in executive function.

Finally, our results show some evidence that increased *PCDHA* and *PCDHG* methylation result in decreased cognitive performance in certain areas in the presence of white matter pathology as indicated via white matter hyperintensity on T2-weighted MRI scans and also increased stroke risk, although further research with more balanced populations will be necessary to confirm this. We would hypothesize that greater synaptogenesis (due to increased *PCDHA* methylation) and increased dendritic arbor loss (due to increased *PCDHG* methylation) result in greater vulnerability to white matter pathology (as more critical neuronal connections are damaged) and also put greater pressure on the aging vasculature (through a mechanism yet to be determined); future research will obviously be necessary to investigate these possibilities further.

This study is subject to several limitations. As the sample is cross-sectional, results are subject to cohort effects, and future longitudinal studies will be necessary to confirm our findings. However, cohort effects are not as strong a potential limitation in our study as we are investigating the effects of interacting variables. Additionally, obtaining brain DNA methylation is infeasible in human studies. While blood DNA methylation has been found to be a reasonable proxy for organ (brain) DNA methylation in animal studies, how well blood DNA methylation reflects brain methylation is unknown in humans. Finally, the PhenoAge metric used to calibrate the DNA clock is only an imperfect measure of biological age and thus leaves open the possibility that there are aging-related variables not completely controlled for.

In sum, our results raise the exciting possibility of retarding normal aging processes via targeted epigenetic modifications (e.g., raising *PCDHA/B* methylation, decreasing *PCDHG* methylation). Epigenetic modifications are much easier to achieve and more affordable than genetic modifications. Methylation at the clustered PCDH loci is found to have a strong moderating effect on both regional brain volume loss and loss of cognitive function during normal aging. Taken together, our results show the potential of epigenetic modifications targeting the normal aging process.

## Supplemental Figures

**Supplemental Figure 1:**
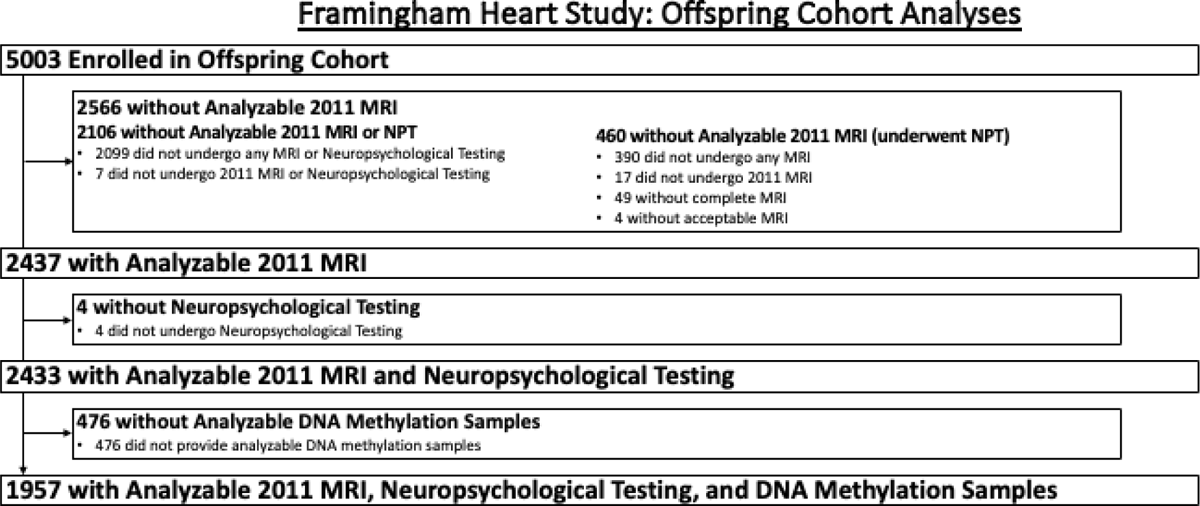
Flow Chart of FHS Subjects and Procedures Included

**Supplemental Figure 2.**
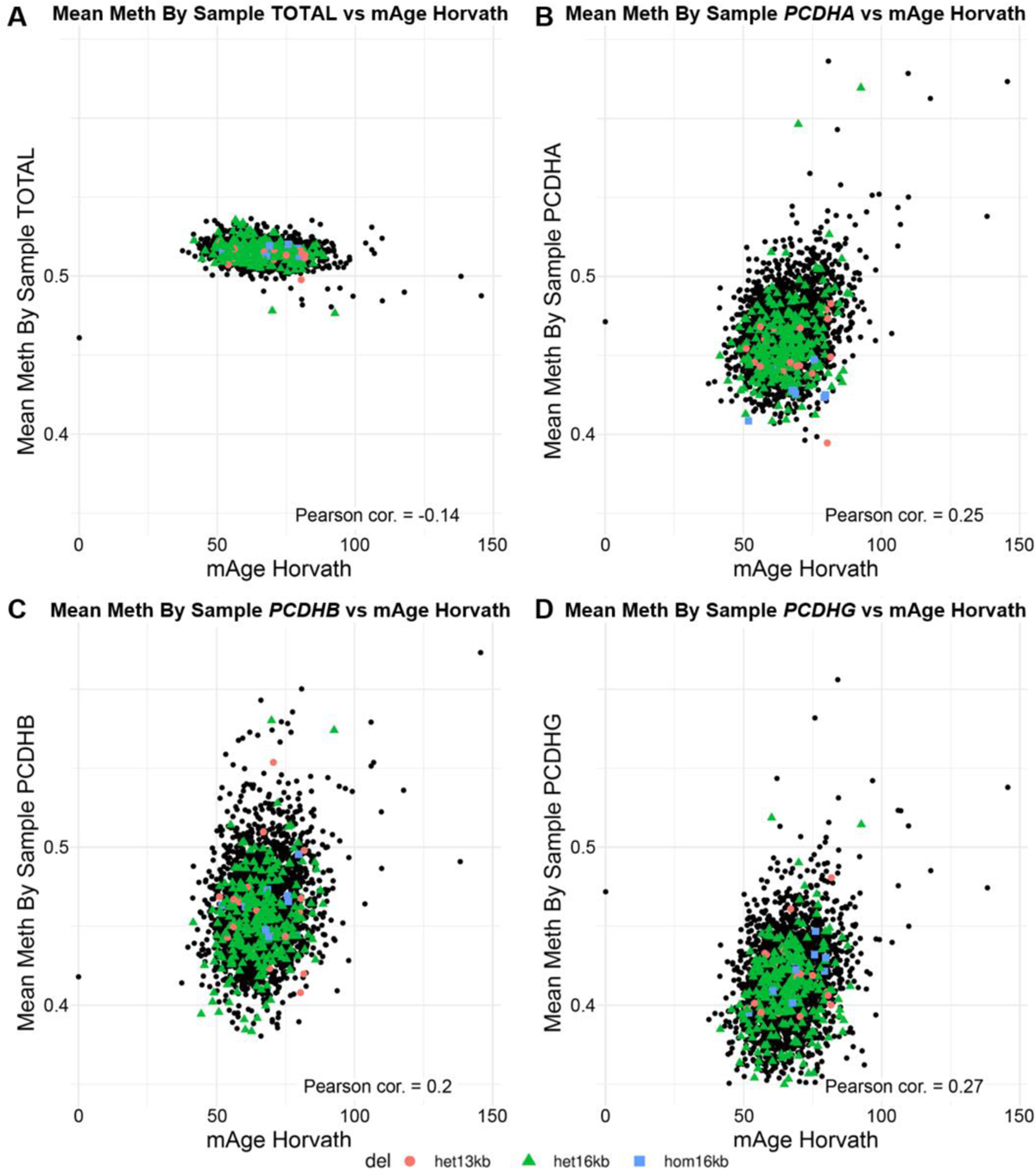
DNA Methylation status of Framingham offspring cohort. Genome-wide mean DNA methylation decreased (A) with increasing age (mAge, Horvath clock), but the three clustered protocadherin loci, *PCDHA* (B), *PCDHB* (C), and *PCDHG* (D) showed marked increase in methylation. Presence of the 13 or 16 kb *PCDHA* delCNVs was associated with decreased mean DNA methylation of the *PCHDA* cluster, with the homozygous delCNV associated with the most severe decrease in DNA methylation. In contrast, the presence of the PCHDA delCNVs had no impact on DNA methylation in the *PCDHB* (C) and *PCDHG* (D) loci.

## Data Availability

All data produced in the present study are available upon reasonable request to the authors.
FHS data were obtained through the database of Genotypes and Phenotypes:

http://dbgap.ncbi.nlm.nih.gov

## References

1 Yagi, T. Clustered protocadherin family. Development, growth & differentiation 50, S131–S140 (2008).

2 Flaherty, E. & Maniatis, T. The role of clustered protocadherins in neurodevelopment and neuropsychiatric diseases. Current opinion in genetics & development 65, 144–150 (2020).

3 Jia, Z. & Wu, Q. Clustered protocadherins emerge as novel susceptibility loci for mental disorders. Frontiers in Neuroscience, 1191 (2020).

4 Peek, S. L., Mah, K. M. & Weiner, J. A. Regulation of neural circuit formation by protocadherins. Cellular and molecular life sciences 74, 4133–4157 (2017).

5 Mountoufaris, G., Canzio, D., Nwakeze, C. L., Chen, W. V. & Maniatis, T. Writing, reading, and translating the clustered protocadherin cell surface recognition code for neural circuit assembly. Annual review of cell and developmental biology 34, 471–493 (2018).

6 McClay, J. L. et al. A methylome-wide study of aging using massively parallel sequencing of the methyl-CpG-enriched genomic fraction from blood in over 700 subjects. Human molecular genetics 23, 1175–1185 (2014).

7 El Hajj, N., Dittrich, M. & Haaf, T. in Seminars in cell & developmental biology. 172–182 (Elsevier).

8 DeCarli, C. et al. Measures of brain morphology and infarction in the framingham heart study: establishing what is normal. Neurobiology of aging 26, 491–510 (2005).

9 Mailman, M. D. et al. The NCBI dbGaP database of genotypes and phenotypes. Nature genetics 39, 1181–1186 (2007).

10 Kohmura, N. et al. Diversity revealed by a novel family of cadherins expressed in neurons at a synaptic complex. Neuron 20, 1137–1151 (1998).

11 Wu, Q. & Maniatis, T. A striking organization of a large family of human neural cadherin-like cell adhesion genes. Cell 97, 779–790 (1999).

12 Chen, W. V. & Maniatis, T. Clustered protocadherins. Development (Cambridge, England) 140, 3297–3302 (2013).

13 Esumi, S. et al. Monoallelic yet combinatorial expression of variable exons of the protocadherin-α gene cluster in single neurons. Nature genetics 37, 171–176 (2005).

14 Hirano, K. et al. Single-neuron diversity generated by Protocadherin-β cluster in mouse central and peripheral nervous systems. Frontiers in molecular neuroscience 5, 90 (2012).

15 Brasch, J. et al. Visualization of clustered protocadherin neuronal self-recognition complexes. Nature 569, 280–283 (2019).

16 Mountoufaris, G. et al. Multicluster Pcdh diversity is required for mouse olfactory neural circuit assembly. *Science (New York*, N.Y.) 356, 411–414 (2017).

17 Lefebvre, J. L., Kostadinov, D., Chen, W. V., Maniatis, T. & Sanes, J. R. Protocadherins mediate dendritic self-avoidance in the mammalian nervous system. Nature 488, 517–521 (2012).

18 Chen, W. V. et al. Pcdhαc2 is required for axonal tiling and assembly of serotonergic circuitries in mice. *Science (New York*, N.Y.) 356, 406–411 (2017).

19 Wang, X., Su, H. & Bradley, A. Molecular mechanisms governing Pcdh-γ gene expression: evidence for a multiple promoter and cis-alternative splicing model. Genes & development 16, 1890–1905 (2002).

20 Hasegawa, S. et al. Distinct and Cooperative Functions for the Protocadherin-α,-β and-γ Clusters in Neuronal Survival and Axon Targeting. Frontiers in molecular neuroscience, 155 (2016).

21 Garrett, A. M. et al. CRISPR/Cas9 interrogation of the mouse Pcdhg gene cluster reveals a crucial isoform-specific role for Pcdhgc4. PLoS genetics 15, e1008554 (2019).

22 Tau, G. Z. & Peterson, B. S. Normal development of brain circuits. Neuropsychopharmacology 35, 147–168 (2010).

23 Weiner, J. A. & Jontes, J. D. Protocadherins, not prototypical: a complex tale of their interactions, expression, and functions. Frontiers in molecular neuroscience 6, 4 (2013).

24 Takeichi, M. The cadherin superfamily in neuronal connections and interactions. Nature Reviews Neuroscience 8, 11–20 (2007).

25 Shapiro, L. & Colman, D. R. The diversity of cadherins and implications for a synaptic adhesive code in the CNS. Neuron 23, 427–430 (1999).

26 Fukuda, E. et al. Down-regulation of protocadherin-α A isoforms in mice changes contextual fear conditioning and spatial working memory. European Journal of Neuroscience 28, 1362–1376 (2008).

27 Garrett, A. M., Schreiner, D., Lobas, M. A. & Weiner, J. A. γ-protocadherins control cortical dendrite arborization by regulating the activity of a FAK/PKC/MARCKS signaling pathway. Neuron 74, 269–276 (2012).

28 Suo, L., Lu, H., Ying, G., Capecchi, M. R. & Wu, Q. Protocadherin clusters and cell adhesion kinase regulate dendrite complexity through Rho GTPase. Journal of molecular cell biology 4, 362–376 (2012).

29 Fan, L. et al. Alpha protocadherins and Pyk2 kinase regulate cortical neuron migration and cytoskeletal dynamics via Rac1 GTPase and WAVE complex in mice. eLife 7, e35242 (2018).

30 Jia, Z. & Wu, Q. Clustered protocadherins emerge as novel susceptibility loci for mental disorders. Frontiers in neuroscience 14, 587819 (2020).

31 Schreiner, D. & Weiner, J. A. Combinatorial homophilic interaction between γ-protocadherin multimers greatly expands the molecular diversity of cell adhesion. Proceedings of the National Academy of Sciences 107, 14893–14898 (2010).

32 Molumby, M. J., Keeler, A. B. & Weiner, J. A. Homophilic protocadherin cell-cell interactions promote dendrite complexity. Cell reports 15, 1037–1050 (2016).

33 Kim, S. et al. Bilingualism for Dementia: Neurological Mechanisms Associated With Functional and Structural Changes in the Brain. Front Neurosci 13, 1224 (2019). 10.3389/fnins.2019.01224

34 Rodríguez-Fornells, A., Cunillera, T., Mestres-Missé, A. & de Diego-Balaguer, R. Neurophysiological mechanisms involved in language learning in adults. Philos Trans R Soc Lond B Biol Sci 364, 3711–3735 (2009). 10.1098/rstb.2009.0130

35 Barak, O. & Tsodyks, M. Working models of working memory. Curr Opin Neurobiol 25, 20–24 (2014). 10.1016/j.conb.2013.10.008

36 De Pitta, M. & Brunel, N. Multiple forms of working memory emerge from synapse-astrocyte interactions in a neuron-glia network model. Proc Natl Acad Sci U S A 119, e2207912119 (2022). 10.1073/pnas.2207912119

37 Fiske, A. & Holmboe, K. Neural substrates of early executive function development. Dev Rev 52, 42–62 (2019). 10.1016/j.dr.2019.100866

38 Constantinidis, C. & Luna, B. Neural Substrates of Inhibitory Control Maturation in Adolescence. Trends Neurosci 42, 604–616 (2019). 10.1016/j.tins.2019.07.004

