## Supplemental Tables for "Complex Regulation of Protocadherin Epigenetics on Aging-Related Brain Health"

Supplemental Table 1. Framingham Heart Study Included vs. Excluded Subject Demographics

| **Characteristics** | **Included (n = 1957)** | **Excluded (n = 3046)** | **Chi-Square /  T-Test p** | **Cohen's D** |
| --- | --- | --- | --- | --- |
| **Males, No. (%)** | 883 (45.12%) | 1541 (50.59%) | 0.0002 | . |
| **Race Reported, No. (%)** | 1942 (99.23%) | 1013 (33.26%) | <0.0001 | . |
| **Reported Race** | . | . | 0.8660 | . |
| **White, No. (%)** | 1924 (99.07%) | 1004 (99.11%) |  |  |
| **African American, No. (%)** | 4 (0.21%) | 2 (0.20%) |  |  |
| **Asian, No. (%)** | 2 (0.10%) | 0 (0.00%) |  |  |
| **Indigenous American, No. (%)** | 1 (0.05%) | 1 (0.10%) |  |  |
| **Multi-Racial, No. (%)** | 11 (0.57%) | 6 (0.59%) |  |  |
| **Ethnicity Reported, No. (%)** | 1758 (89.83%) | 905 (29.71%) | <0.0001 | . |
| **Hispanic or Latino, No. (%)** | 5 (0.28%) | 8 (0.88%) | 0.0704 | . |
| **Education Reported, No. (%)** | 1956 (99.95%) | 928 (30.47%) | <0.0001 | . |
| **Education Level** | . | . | <0.0001 | . |
| **High School Degee, No. (%)** | 651 (33.28%) | 382 (41.16%) |  |  |
| **Some College, No. (%)** | 497 (25.41%) | 256 (27.59%) |  |  |
| **College Graduate, No. (%)** | 808 (41.31%) | 290 (31.25%) |  |  |
| **History of Stroke, No. (%)** | 98 (5.01%) | 264 (8.67%) | <0.0001 | . |
| **Stroke Type** | . | . | 0.8776 | . |
| **Atherothrombotic Infarction of Brain, No. (%)** | 58 (59.18%) | 151 (57.20%) |  |  |
| **Cerebral Embolism, No. (%)** | 22 (22.45%) | 67 (25.38%) |  |  |
| **Cerebral Vascular Accident, No. (%)** | 2 (2.04%) | 6 (2.27%) |  |  |
| **Intracerebral Hemorrhage, No. (%)** | 12 (12.24%) | 25 (9.47%) |  |  |
| **Subarachnoid Hemorrhage, No. (%)** | 4 (4.08%) | 15 (5.68%) |  |  |
| **Age at Neuropsychological Testing** |  |  | <0.0001 | 0.5080 |
| **No.** | 1957 | 936 |  |  |
| **mean (SD)** | 60.87 (9.05) | 65.80 (10.32) |  |  |
| **range** | 34 - 88 | 36 - 93 |  |  |
| **median (Q1, Q3)** | 60 (54, 68) | 66 (58, 73.25) |  |  |
| **WRAT-3 Reading** |  |  | 0.0006 | 0.1393 |
| **No.** | 1933 | 895 |  |  |
| **mean (SD)** | 48.81 (5.20) | 48.08 (5.23) |  |  |
| **range** | 15 - 57 | 19 - 57 |  |  |
| **median (Q1, Q3)** | 49 (46, 53) | 48 (45, 52) |  |  |
| **WAIS Similarities** |  |  | <0.0001 | 0.4030 |
| **No.** | 1954 | 927 |  |  |
| **mean (SD)** | 16.99 (3.55) | 15.36 (4.48) |  |  |
| **range** | 2 - 25 | 0 - 25 |  |  |
| **median (Q1, Q3)** | 17 (15, 19) | 16 (13, 18) |  |  |

Notes: Cerebral Vascular Accident includes definite, questionable, and other types. WRAT-3=Wide Range Achievement Test, 3rd Edition. WAIS=Wechsler Adult Intelligence Scale.

Supplemental Table 2. Association of Between PCDH Loci (alpha, beta, gamma) and Total DNA Methylation Levels and Chronological Age-Related Decline in Regional Brain Volumes

| ^Brain Volume^ | ^Accelerated^  ^Age^ | ^Age^ | ^Age x Accelerated Age^ | ^PCDHA^ | ^Age x PCDHA^ | ^PCDHA2^ | ^PCDHB^ | ^Age x PCDHB^ | ^PCDHB2^ | ^PCDHG^ | ^Age x PCDHG^ | ^PCDHG2^ | ^Total^ | ^Age x Total^ | ^Total2^ |
| --- | --- | --- | --- | --- | --- | --- | --- | --- | --- | --- | --- | --- | --- | --- | --- |
| ^Temporal Lobe White Matter^ | ^0.010 (0.031), 0.7457^ | ^-0.377 (0.023), 0.0000^ | ^0.007 (0.003), 0.0371^ | ^-0.816 (0.265), 0.0021*^ | ^0.099 (0.027), 0.0003*^ | ^-0.133 (0.088), 0.1302^ | ^-^ | ^-^ | ^-^ | ^0.456 (0.257), 0.0762^ | ^-0.063 (0.027), 0.0202†^ | ^-^ |  |  |  |
| ^Temporal Lobe Gray Matter^ | ^-0.035 (0.038), 0.3629^ | ^-0.385 (0.029), 0.0000^ | ^0.009 (0.004), 0.0329^ | ^-0.606 (0.259), 0.0194†^ | ^0.084 (0.027), 0.0021*^ | ^-^ | ^-^ | ^-^ | ^-^ | ^-^ | ^-^ | ^-^ | ^-^ | ^-^ | ^-^ |
| ^Parietal^  ^White Matter^ | ^0.031 (0.038), 0.4127^ | ^-0.328 (0.029), 0.0000^ | ^0.005 (0.004), 0.2574^ | ^-0.809 (0.324), 0.0126†^ | ^0.134 (0.034), 0.0001*^ | ^-0.264 (0.108), 0.0143^ | ^-^ | ^-^ | ^-^ | ^0.884 (0.315), 0.0050*^ | ^-0.079 (0.033), 0.0172†^ | ^-^ | ^-^ | ^-^ | ^-^ |
| ^Parietal^  ^Gray Matter^ | ^-0.086 (0.034), 0.0108^ | ^-0.310 (0.026), 0.0000^ | ^0.006 (0.004), 0.0844^ | ^-0.674 (0.282), 0.0170†^ | ^0.071 (0.029), 0.0142†^ | ^-^ | ^-^ | ^-^ | ^-^ | ^0.541 (0.282), 0.0551^ | ^-0.052 (0.030), 0.0840^ | ^-^ | ^-^ | ^-^ | ^-^ |
| ^Frontal Lobe White Matter^ | ^0.047 (0.070), 0.5036^ | ^-0.740 (0.053), 0.0000^ | ^0.011 (0.008), 0.1552^ | ^-0.731 (0.495), 0.1400^ | ^0.138 (0.053), 0.0090*^ | ^-0.315 (0.201), 0.1173^ | ^-^ | ^-^ | ^-^ | ^-^ | ^-^ | ^-^ | ^-^ | ^-^ | ^-^ |
| ^Frontal Lobe Gray Matter^ | ^-0.089 (0.057), 0.1158^ | ^-0.663 (0.043), 0.0000^ | ^0.010 (0.006), 0.1027^ | ^-0.932 (0.479), 0.0516^ | ^-^ | ^-^ | ^0.227 (0.477), 0.6348^ | ^0.112 (0.042), 0.0069*^ | ^-^ | ^-^ | ^-^ | ^-^ | ^-^ | ^-^ | ^-^ |
| ^Occipital White Matter^ | ^0.009 (0.024), 0.7103^ | ^-0.146 (0.018), 0.0000^ | ^0.007 (0.003), 0.0066^ | ^-0.787 (0.198), 0.0001*^ | ^0.044 (0.017), 0.0101†^ | ^-^ | ^-^ | ^-^ | ^-^ | ^0.653 (0.201), 0.0012*^ | ^-^ | ^-0.132 (0.074), 0.0760^ | ^-^ | ^-^ | ^-^ |
| ^Occipital Gray Matter^ | ^-0.049 (0.025), 0.0520^ | ^-0.259 (0.019), 0.0000^ | ^0.006 (0.003), 0.0423^ | ^-0.764 (0.213), 0.0003*^ | ^0.050 (0.018), 0.0057*^ | ^-^ | ^-^ | ^-^ | ^-^ | ^0.461 (0.212), 0.0295†^ | ^-^ | ^-^ | ^-^ | ^-^ | ^-^ |

*Red

†Yellow

Abbreviations:

protocadherin alpha (PCHDA)

protocadherin beta (PCDHB)

protocadherin gamma (PCDHG)

PCDHA^2.^  =quadrant term in the model

Supplemental Table 3. Association of Between PCDH Loci (alpha, beta, gamma) DNA Methylation Levels and Chronological Age-Related Decline in Neurocognitive Outcomes

| Cognitive Outcome | Accelerated Age | Age | Age x Accelerated Age | PCDHA | Age x PCDHA | PCDHA^2^ | PCDHB | Age x PCDHB | PCDHB^2^ | PCDHG | Age x PCDHG | PCDHG^2^ | Total | Age x Total | Total^2^ |
| --- | --- | --- | --- | --- | --- | --- | --- | --- | --- | --- | --- | --- | --- | --- | --- |
| LMI | -1.351 (1.173), 0.2497 | -6.648 (0.876), 0.0000 | 0.281 (0.131), 0.0322 | - | - | - | - | - | - | - | - | - | - | - | - |
| LMD | -2.602 (1.241), 0.0362 | -8.375 (0.927), 0.0000 | 0.288 (0.139), 0.0384 | - | - | - | - | - | - | - | - | - | - | - | - |
| LMR | -0.031 (0.425), 0.9423 | -1.385 (0.317), 0.0000 | 0.054 (0.048), 0.2577 | - | - | - | - | - | - | - | - | - | - | - | - |
| VRI | -2.310 (1.036), 0.0260 | -12.393 (0.774), 0.0000 | 0.135 (0.116), 0.2449 | - | - | - | - | - | - | - | - | - | - | - | - |
| VRD | -1.705 (1.090), 0.1178 | -13.089 (0.814), 0.0000 | 0.124 (0.122), 0.3079 | - | - | - | - | - | - | - | - | - | - | - | - |
| VRR | -0.572 (0.339), 0.0916 | -2.916 (0.253), 0.0000 | 0.031 (0.038), 0.4198 | - | - | - | - | - | - | - | - | - | - | - | - |
| PASI | -1.357 (1.115), 0.2237 | -10.001 (0.835), 0.0000 | 0.132 (0.124), 0.2894 | - | - | - | - | - | - | - | - | - | -13.048 (7.300), 0.0740 | - | - |
| PASD | -0.198 (0.488), 0.6857 | -4.231 (0.374), 0.0000 | 0.032 (0.054), 0.5586 | - | - | - | - | - | - | -4.830 (3.287), 0.1418 | - | - | - | - | - |
| BNT30 | -1.689 (0.829), 0.0418 | -7.655 (0.619), 0.0000 | 0.029 (0.093), 0.7554 | - | - | - | - | - | - | - | - | - | - | - | - |
| WRAT | -6.023 (1.824), 0.0010 | -8.591 (1.406), 0.0000 | 0.405 (0.203), 0.0461 | -8.735 (15.356), 0.5695 | 3.984 (1.587), 0.0121† | - | - | - | - | -5.679 (15.380), 0.7120 | -4.526 (1.641), 0.0059* | - | - | - | - |
| SIM | -3.695 (1.196), 0.0020 | -9.670 (0.918), 0.0000 | 0.152 (0.133), 0.2542 | - | - | - | -6.600 (10.342), 0.5234 | 3.414 (1.131), 0.0026* | - | 4.840 (10.587), 0.6476 | -2.712 (1.157), 0.0192† | - | - | - | - |
| HVOT | -0.390 (0.974), 0.6891 | -11.588 (0.727), 0.0000 | 0.247 (0.109), 0.0232 | - | - | - | - | - | - | - | - | - | - | - | - |
| TRA | 0.258 (0.065), 0.0001 | 0.758 (0.050), 0.0000 | 0.003 (0.007), 0.6825 | -0.346 (0.465), 0.4570 | 0.105 (0.051), 0.0393† | -0.346 (0.190), 0.0680 | - | - | - | - | - | - | - | - | - |
| TRB | 0.470 (0.306), 0.1245 | 2.982 (0.228), 0.0000 | 0.024 (0.034), 0.4878 | - | - | - | - | - | - | - | - | - | 3.972 (2.111), 0.0600 | 0.654 (0.240), 0.0066* | 1.449 (0.737), 0.0493 |

*Red

†Yellow

Abbreviations:

WMS Logical Memory Immediate Recall (LMI)

WMS Logical Memory Delayed Recall (LMD)

WMS Logical Memory Recognition (LMR)

WMS Visual Reproductions Immediate Recall (VRI)

WMS Visual Reproductions Delayed Recall (VRD)

WMS Visual Reproductions Recognition (VRR)

WMS Verbal Paired Associates Total Immediate Recall (PASI)

WMS Verbal Paired Associates Total Delayed Recall (PASD)

Boston Naming Test (BNT30)

Wide Range Achievement Test (WRAT) Word Reading

WAIS-IV Similarities (SIM)

Hooper Visual Organization Test (HVOT)

Trail Making Test Part A (TRA)

Trail Making Test Part B (TRB)

protocadherin alpha (PCHDA)

protocadherin beta (PCDHB)

protocadherin gamma (PCDHG)

PCDHA^2^ = quadrant term in the model

Supplemental Table 4. Association of Between PCDH Loci (alpha, beta, gamma) and Total DNA Methylation Levels, White Matter Hyperintensity Volume, and Cognitive Outcomes

| Cognitive Outcome | White Matter Hyper-intensities  Volume | PCDHA | White Matter Hyper-intensities  Volume x PCDHA | PCDHA^2^ | PCDHB | White Matter Hyper-intensities  Volume x PCDHB | PCDHB^2^ | PCDHG | White Matter Hyper-intensities  Volume x PCDHG | PCDHG^2^ | Total | White Matter Hyper-intensities  Volume x Total | Total^2^ |
| --- | --- | --- | --- | --- | --- | --- | --- | --- | --- | --- | --- | --- | --- |
| LMI | 9.981 (8.417), 0.2358 | - | - | - | - | - | - | - | - | - | - | - | - |
| LMD | 4.359 (8.908), 0.6246 | - | - | - | - | - | - | - | - | - | - | - | - |
| LMR | -1.841 (3.045), 0.5454 | - | - | - | - | - | - | - | - | - | - | - | - |
| VRI | -10.621 (7.443), 0.1537 | - | - | - | - | - | - | - | - | - | - | - | - |
| VRD | -12.327 (7.823), 0.1153 | - | - | - | - | - | - | - | - | - | - | - | - |
| VRR | -0.027 (2.433), 0.9912 | - | - | - | - | - | - | - | - | - | - | - | - |
| PASI | -1.200 (7.963), 0.8802 | - | - | - | - | - | - | - | - | - | -12.746 (7.303), 0.0811 | - | - |
| PASD | -2.122 (3.471), 0.5411 | - | - | - | - | - | - | -4.650 (3.287), 0.1573 | - | - | - | - | - |
| BNT30 | 5.188 (6.102), 0.3953 | -7.438 (7.127), 0.2968 | -7.395 (2.757), 0.0074* | - | 8.482 (7.044), 0.2287 | 5.120 (2.756), 0.0634 | - | - | - | - | - | - | - |
| WRAT | 21.823 (13.223), 0.0990 | - | - | - | 4.351 (15.714), 0.7819 | 15.137 (7.314), 0.0386† | - | -14.113 (16.030), 0.3787 | -19.172 (7.457), 0.0102† | - | - | - | - |
| SIM | 6.712 (8.688), 0.4399 | -2.351 (8.040), 0.7700 | -8.816 (3.483), 0.0115† | - | - | - | - | - | - | - | - | - | - |
| HVOT | -12.776 (7.020), 0.0689 | - | - | - | - | - | - | - | - | - | - | - | - |
| TRA | 1.025 (0.465), 0.0276 | - | - | - | - | - | - | - | - | - | - | - | - |
| TRB | 11.782 (2.168), 0.0000 | - | - | - | - | - | - | - | - | - | 2.971 (1.981), 0.1340 | 4.377 (1.199), 0.0003* | - |

*Red

†Yellow

Abbreviations:

WMS Logical Memory Immediate Recall (LMI)

WMS Logical Memory Delayed Recall (LMD)

WMS Logical Memory Recognition (LMR)

WMS Visual Reproductions Immediate Recall (VRI)

WMS Visual Reproductions Delayed Recall (VRD)

WMS Visual Reproductions Recognition (VRR)

WMS Verbal Paired Associates Total Immediate Recall (PASI)

WMS Verbal Paired Associates Total Delayed Recall (PASD)

Boston Naming Test (BNT30)

Wide Range Achievement Test (WRAT) Word Reading

WAIS-IV Similarities (SIM)

Hooper Visual Organization Test (HVOT)

Trail Making Test Part A (TRA)

Trail Making Test Part B (TRB)

protocadherin alpha (PCHDA)

protocadherin beta (PCDHB)

protocadherin gamma (PCDHG)

PCDHA^2.^  = quadrant term in the model

Supplemental Table 5. Clinical Risk

| Clinical Outcome | Accelerated Age | Age | Age x Accelerated Age | PCDHA | Age x PCDHA | PCDHA^2^ | PCDHB | Age x PCDHB | PCDHB^2^ | PCDHG | Age x PCDHG | PCDHG^2^ | Total | Age x Total | Total^2^ |
| --- | --- | --- | --- | --- | --- | --- | --- | --- | --- | --- | --- | --- | --- | --- | --- |
| Stroke | 0.020 (0.018), 0.2818 | 0.058 (0.013), 0.0000 | 0.000 (0.002), 0.8744 | 0.210 (0.130), 0.1068 | - | - | - | - | - | 0.064 (0.032), 0.0483† | - | -1.087 (0.385), 0.0048 | - | - | - |
| MCI | 0.020 (0.024), 0.4087 | 0.168 (0.015), 0.0000 | 0.002 (0.002), 0.4253 | - | - | - | - | - | - | - | - | - | 0.065 (0.027), 0.0170† | - | -3.924 (0.466), 0.0000 |
| Alzheimer | 0.042 (0.038), 0.2788 | 0.183 (0.022), 0.0000 | -0.002 (0.003), 0.5451 | - | - | - | - | - | - | - | - | - | -5.539 (0.719), 0.0000* | - | - |

*Red

†Yellow

Abbreviations:

Mild Cognitive Impairment (MCI)

protocadherin alpha (PCHDA)

protocadherin beta (PCDHB)

protocadherin gamma (PCDHG)

PCDHA^2.^  = quadrant term in the model
